# Predicting Disease Progression in COVID19: A Score Based On Lab Tests At Time Of Diagnosis

**DOI:** 10.1101/2020.05.05.20088906

**Authors:** Mohammad H. Jamal, Suhail A. Doi, Sarah AlYouha, Sulaiman Almazeedi, Mohannad Al-Haddad, Ali Al-Muhaini, Fahad Al-Ghimlas, Salman Al-Sabah

## Abstract

**Background:** COVID19 is worldwide pandemic that is mild in the majority of patients but can result in a pneumonia like illness with progression to acute respiratory distress syndrome and death. Predicting the disease severity at time of diagnosis can be helpful in prioritizing hospital admission and resources.

**Methods:** We prospectively recruited 1096 consecutive patients with COVID19 from the Jaber Hospital, a COVID19 facility in Kuwait, between 24 February and 20 April 2020. The primary endpoint of interest was disease severity defined algorithmically. Predefined risk variables were collected at the time of PCR based diagnosis of the infection. Prognostic model development used 5-fold cross-validated regularized logit regression. The cohort was divided into a training and validation cohort and all model development proceeded on the training cohort.

**Results:** There were 643 patients with clinical course data of whom 94 developed severe COVID19. In the final model, age, CRP, procalcitonin, lymphocyte and monocyte percentages and serum albumin were independent predictors of a more severe illness course. The final prognostic model demonstrated good discrimination, calibration and internal validity.

**Conclusion:** We developed and validated a simple score calculated at time of diagnosis that can predict patients with severe COVID19 disease.

## Introduction

The outbreak of pneumonia in the Hubei province of the People’s Republic of China in December 2019 was identified to be due to a novel corona virus, namely severe acute respiratory syndrome coronavirus 2 (SARS-CoV-2). The disease termed COVID19 became a pandemic affecting more than 3,419,000 people worldwide with around 243,000 deaths (up to 2 May 2020).^1^ The majority of patients with COVID19 recover, but a subset of patients develop severe disease characterized by a cytokine storm that increases the risk of mortality.^2^ The main cause of mortality in those patients is acute respiratory distress syndrome (ARDS) or septic shock which occurs in 15-20% of patients.^3^

A study on the New York experience on hospitalized patients with COVID19 reported that out of 5700 patients, 373 (14.2%) were treated in the ICU, 320 (12.2%) received invasive mechanical ventilation, 81 (3.2%) were treated with kidney replacement therapy, and 553 (21%) died.^4^ In many countries around the world admission to hospitals is reserved for those with severe symptoms which usually do not develop from the onset of symptoms or from the time of diagnosis with the PCR test. Patients with severe symptoms present usually after a mild first phase of the disease.^5^

To date there has not been an effective therapeutic modality in the form of an antiviral medication or vaccine against the disease, but many regimens have been tested and some experts suggested suppressing the immune system to avoid the cytokine storm leading to ARDS.^2^ All these proposed treatments carry their own risks, and immunosuppression might increase the risks for other viral and bacterial infections thus hardly justifying their use in mildly symptomatic patients.

The challenge today is determining and stratifying which patient is likely to progress to severe disease at time of diagnosis. Answering this question might justify early treatment and admission to hospitals. In this study we examine the initial cohort of patients in Kuwait to determine what risk factors at time of positivity of a test can predict a worse outcome, as all patients with a positive test even if asymptomatic are admitted to a single center in the State of Kuwait.

## METHODS

### Study Design

We obtained the ethical approval from the Kuwait Ministry of Health ethical review committee. We carried out a prospective cohort study to explore the factors associated with COVID19 severity amongst COVID19 in-patients admitted to the Infectious Diseases Hospital in Kuwait between 24 February 2020 and 20 April 2020.

### Inclusion Criteria and Case Definition

All consecutive patients meeting the case definition and who had tested positive are diagnosed to have COVID19 and admitted for quarantine and observation. Testing for COVID19 was undertaken via real-time reverse-transcriptase–polymerase-chain-reaction (RT-PCR) assay of nasal swab specimens. All diagnostic tests were performed at the Jaber Hospital laboratory. All positive patients stay in hospital till they have had resolution of symptoms (afebrile for more than 72 hours plus saturation ≥ 94%) so long as they are also more than 7 days since symptoms onset, have completed 14 days since testing positive and are Improving or have regression of abnormalities on imaging. Discharge occurs after two consecutive negative tests >24h apart. A standardized form was completed prospectively for data collection, including demographic data, clinical data and radiographic / laboratory results.

### Severity grouping (main outcome)

We pre-specified the main outcome to be moderate-severe COVID19 defined based on need for hospital support, while mild cases will have a mild clinical course needing only symptomatic management (the majority). The severity grouping algorithm was determined prospectively as follows:

1. Assign missing status to everyone
2. Assign moderate to severe status to those with hospital course that led to:
  a. Death
  b. Consolidation on chest x-ray or shortness of breath on admission
  c. ICU admission
  d. Hospital stay >14 days AND in hospital receiving active treatment
3. Assign moderate to severe status additionally to those receiving the following treatments regardless of hospital stay or discharge status:
  a. Systemic gluococorticoids
  b. Intravenous immunoglobulin
  c. Oxygen therapy
  d. Non-invasive ventilation OR mechanical ventilation
  e. Extracorporeal membrane oxygenation
  f. Continuous renal replacement therapy
4. Assign mild status to those meeting the following:
  a. Not on treatment, not discharged and duration in hospital >10 days
  b. Discharged and no treatment received
  c. Discharged and duration in hospital >14 days and severity score not already assigned to severe status above

Apart from the severity group outcome, ICU admission and death were defined as outcomes for the purposes of model evaluation but not in model building as they were subsumed within the main outcome.

### Statistical Analysis

Categorical variables were summarized using percentages while continuous variables were summarized using medians with interquartile ranges. Potential predictors for the occurrence of severe COVID19 were investigated using regularized logistic regression (a machine learning algorithm).

All promising predictor variables (demographics, laboratory test results, comorbidities and selected symptoms; see details in supplementary material with details of tests used and the variables considered) with a plausible biological reason for inclusion were assessed at admission prior to the outcome being known. Variables used in the outcome definition were excluded from condideartion as a predictor. All predictors included in the analysis were converted to variable scores (if a continuous variable) prior to entry into the regularized regression procedure. The scores for these transformed continuous variables were 0 or 1 representing values below or above the median while for untransformed binary variables was also 0 or 1 (absent vs present respectively). We decided to categorize continuous variables for the often criticized goal of aiding clinical interpretation and maintaining simplicity. While this may have introduced loss of information we did not plan to reconsider this approach unless there was a problem with predictive performance since using continuous variables would make the model less applicable to rapid implementation during the pandemic. Of note cut-points for continuous variables were predefined (at the median) and decided upon prior to data analysis.

The regularized regression procedure used was a 5-fold cross validated lasso logit regression and we identified the best model using the largest value of the tuning parameter that was within one standard deviation of the optimum value (i.e. the value that minimizes the mean squared error of prediction), which leads to a more parsimonious model. Then an unrestricted logit model was fitted to the selected set of predictors from the lasso model and these coefficients used to determine a preliminary severity risk score for COVID19. This was done by rounding the beta coefficient from the unrestricted logit model on the selected variables to use as integer weights for each variable score. The assigned weights multiplied by variable score were then summed to compute that patient’s severity score (Table 2). Regularized regression was run using lassopack in Stata.^6^ Time to event analyses were not considered even though this was a dynamic cohort because duration of stay varied based on physician discretion and there was no real risk of over-representing those with the moderate to severe disease outcome for COVID19.

To assess discrimination of the model, the C statistic (area under the receiver operating characteristic curve) and its 95% CI were computed. To test model internal validity, a straightforward and fairly popular approach was used which was to randomly split the data in two parts: one to develop the model and another to measure its performance. We used randomtag in Stata to tag the two data-sets (N=700 for training and the rest for validation) and the score was developed on the training data-set. Operating characteristics of the score were then assessed using application to the full data-set. Finally, calibration of the model was assessed using pmcalplot in Stata.^7^ All analyses were performed using Stata MP version 15 (College Station, TX, USA) and the confidence level was set at 95%.

## RESULTS

### Baseline Characteristics

From 24 February 2020 till 20 April 2020, 1096 consecutive patients with COVID19 were included in the study. 888 (81%) were males and 335 (30%) had comorbid conditions. The mean age was 41.4 years (range, 1-93 years) and the average time between the onset of symptoms and admission was 4.2 days (95% CI 3.8 - 4.5 days). The most frequent clinical symptoms of COVID19 were chills and cough. Of the 1096 patients, clinical course could be defined (see next section) for 643 patients (the rest had not yet reached the outcome) and these formed the cohort for the model development. Details of the basic characteristics of these patients are given in Table 1. Of note, the final severity score could be computed for 1008 individuals (642/700 in the training cohort and 366/396 in the validation cohort).

**Table 1:** Baseline Characteristics of patients with COVID-19

| Characteristic | Median (IQR) or<br>N (%)<br>With clinical<br>course data<br>(N=643) | Median (IQR)<br>or<br>N (%)<br>Training cohort<br>(N=700) | Median (IQR) or<br>N (%)<br>Validation<br>cohort (N=396) | Median (IQR)<br>or<br>N (%)<br>All data<br>(N=1096) |
| --- | --- | --- | --- | --- |
| Age | 39 (29 - 54) | 40 (31 - 51) | 39 (31 - 52) | 40 (31 - 51) |
| Males | 466 (72.5%) | 572 (81.7%) | 316 (79.8%) | 888 (81.0%) |
| Nationality |  |  |  |  |
| Asian* | 269 (40.4%) | 400 (57.1%) | 217 (54.8) | 617 (56.3%) |
| Kuwaiti | 257 (40.0%) | 179 (25.6%) | 118 (29.8) | 297 (27.1%) |
| Others | 126 (19.6%) | 121 (17.3%) | 61 (15.4) | 182 (16.6%) |
| Duration of stay<br>(days)** | 17 (8 - 23) | 9 (3 - 17) | 10 (3 - 18) | 9 (3 - 18) |
| ICU admission | 42 (6.5%) | 27 (3.9%) | 15 (3.8%) | 42 (3.8%) |
| Death | 19 (2.9%) | 10 (1.4%) | 9 (2.3%) | 19 (1.7%) |

\*India/Bangladesh/Philippines
\*\* Shorter stay for the whole cohort as included those more recently admitted whose course was yet to be determined

The clinical course was a mild COVID19 course for 549 patients, a moderate – severe COVID19 course for 94 patients leading to 15 events per variable for the selected model. The course of illness was set to missing for 453 patients. Of the 94 severe cases, 42 were admitted to the intensive care unit for acute respiratory care and 19 died.

In total, 581 out of 643 patients had data on both the clinical course and the severity score parameters and of these 363 were randomly allocated to the training set and 218 to the validation set (using randomtag in Stata). The prediction model was built on the 363 subjects in the training data-set and validated on the 218 in the validation data-set. However final severity scores were calculable on 1008 patients and these were used for score assessment when the outcome was not clinical course in hospital.

### Severity Score of COVID19

Variables selected through lasso logit regression included age, CRP, procalcitonin, lymphocyte percentage, monocyte percentage and serum albumin (Table 2). The range of scores seen were from minus 32 to plus 22 for this cohort.

**Table 2:** Variable scores used in the multivariable logistic regression model, ranges of variables by the variable score and weights used for scoring

| Variable | Score |  | Weight | Example of a severity score for a patient that died |
| --- | --- | --- | --- | --- |
|  | 0 | 1 |  |  |
|  | Values |  |  |  |
| Age (years) | <=40 | 41+ | 4 | 1x4=4 |
| C-reactive protein (mg/L) | <=6 | 7+ | 2 | 1x2=2 |
| Procalcitonin (ng/ml) | <=0.049 | 0.05+ | 16 | 1x16=16 |
| Lymphocyte% | <=31.4 | 31.5+ | -9 | 0x-9=0 |
| Monocyte% | <=9.1 | 9.2+ | -8 | 0x-8=0 |
| Albumin (g/L) | <=39.4 | 39.5) | -15 | 0x-15=0 |
|  |  |  |  | Total=22 |

\*possible range -32 to 22
\*\*Low risk <=-7; high risk >=16

The area under the curve (AUC) for the training sample (N=363) model was equal to 0.834 (95% CI, 0.779-0.889), which indicates good model discrimination (figure 1). The validation cohort (N=218) demonstrated equally good discrimination with AUC 0.794 (95% CI, 0.710 – 0.879) and is also depicted in Figure 1. A calibration plot of observed against expected probabilities for assessment of prediction model performance on the validation cohort demonstrated reasonably good model calibration (Supplementary material Figure A). As expected, the training cohort showed perfect calibration (Supplementary material Figure B)..

**Figure 1:**
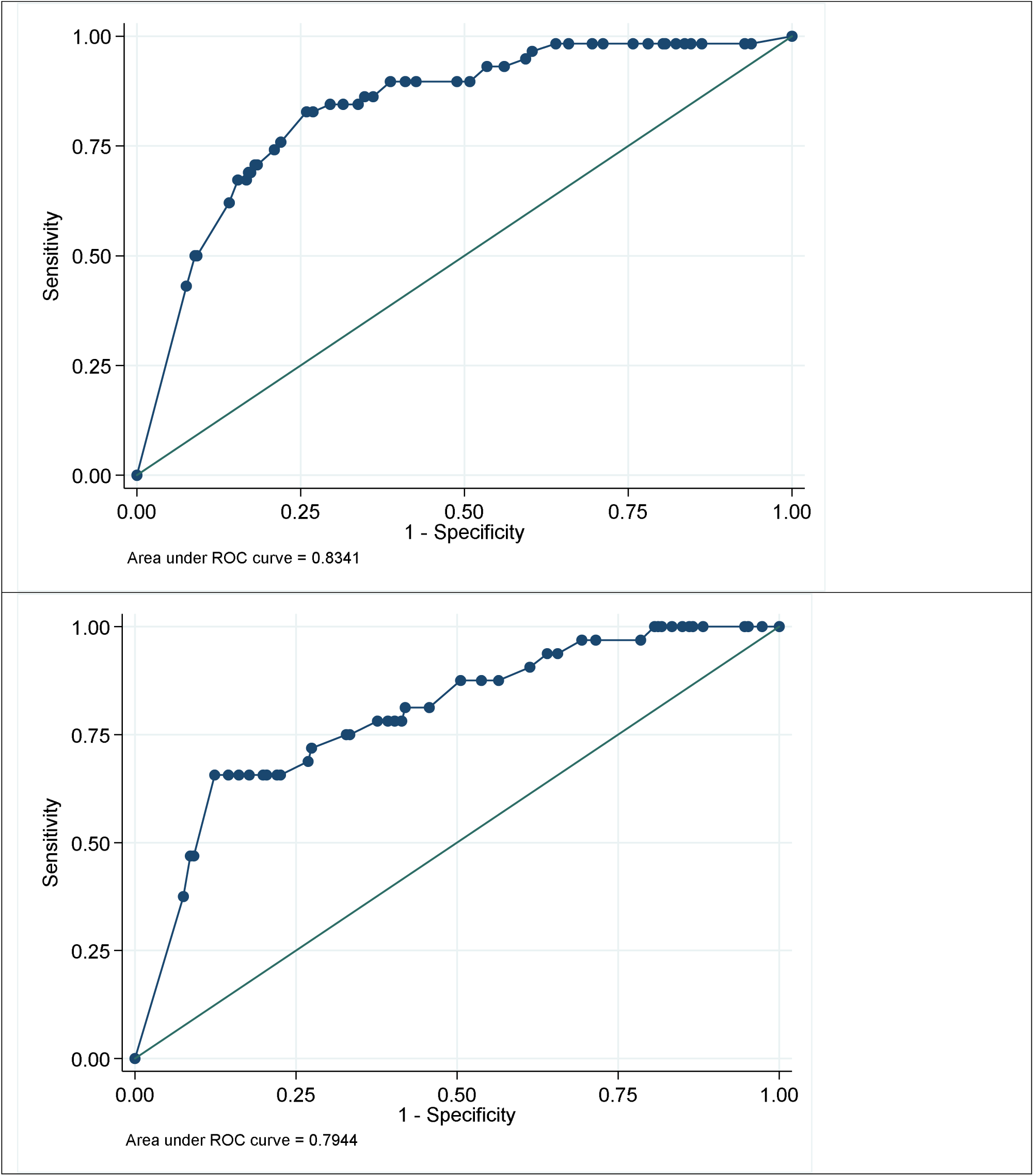
Area under the receiver operating characteristic curve for the severity score in the training (top) and validation (bottom) cohorts

A cutoff for low, intermediate and high risk was chosen according to the score’s performance (thresholds at 90% sensitivity and 90% specificity) and patients with a score −7 or less were at low risk and those with a score 16 or above were at high risk of more severe illness requiring hospital management. The risk groups also demonstrated good discrimination of the various outcomes (Table 3) and the interval likelihood ratios by risk level (based on the score) are presented in Table 4. From the interval likelihood ratios and the baseline prevalence of a severe clinical course (14.6%) we could compute posterior probabilities. The interval likelihood ratio for the severe risk level was 5.33. This means that for a patient at this risk level, the posterior probability of a severe clinical course is 48%. For a patient at the low risk level (with a score of - 7 or less), the posterior probability of a severe clinical course is down to only 4%. Given the way the thresholds were created, at the low risk threshold the score has a sensitivity of 90 % and similarly at the high risk threshold it has a specificity of 90%.

**Table 3:** Odds ratio of outcomes by category of severity score

| Outcome | Severity score | Risk level | Odds ratio | 95% CI |
| --- | --- | --- | --- | --- |
| Moderate to severe course in hospital | <= -7 | Low | 1 (reference) |  |
|  | -6 to 15 | Intermediate | 4.27 | 2.07 - 8.82 |
|  | >=16 | High | 23.66 | 11.10 - 50.43 |
| ICU admission | <= -7 | Low | 1 (reference) |  |
|  | -6 to 15 | Intermediate | 17.79 | 2.32 - 136.57 |
|  | >=16 | High | 159.80 | 21.43 - 1191.42 |
| Death | <= -7 | Low | 1 (reference) |  |
|  | -6 to 15 | Intermediate | 5.34 | 0.59 - 48.00 |
|  | >=16 | High | 66.17 | 8.56 - 511.55 |

**Table 4:** Interval likelihood ratios by risk level

| Risk Level | Mod-Severe<sup>a</sup> | Mild<sup>b</sup> | Likelihood ratio | 95% CI |
| --- | --- | --- | --- | --- |
| Low | 10 | 242 | 0.225 | 0.125 to 0.407 |
| Intermediate | 36 | 204 | 0.963 | 0.732 to 1.266 |
| High | 44 | 45 | 5.334 | 3.761 to 7.566 |
| Total | 90 | 491 |  |  |

<sup>a</sup> composite = 1
<sup>b</sup> composite = 0

## Discussion

In this single center prospective study using a machine learning algorithm we report a prognostic score that stratifies patients with COVID19 according to the risks of severe illness (clinically and radiologically), ICU admission or death based on age and laboratory tests at presentation. We have found that higher age, higher CRP, higher procalcitonin, lower lymphocyte percentage, lower monocyte percentage and lower serum albumin were the most significant predictors of progression of disease, the need for medical support and treatment and the need for ICU admission or death if the score put the patient in the high risk category. The AUC of the model was equal to 0.83 (95% CI, 0.779-0.889), which indicates good discrimination between the groups. It is important to note that this cohort of patients include all the patients who tested PCR positive for COVID19 in Kuwait, even if they were asymptomatic, and all these patients underwent the laboratory investigations tested in this model at time of diagnosis.

Gong et al,^8^ created a COVID19 severity model based on lab tests of 372 non severe patients who were admitted to three clinical centers in Wuhan. They found that old age, and higher serum lactate dehydrogenase, C-reactive protein, the red blood cell distribution width, blood urea nitrogen, direct bilirubin and lower albumin, are associated with severe COVID19. Our initial analysis using standard regression techniques also picked up these same variables (except lactate dehydrogenase) but depending on the randomly selected training samples the selection process remained very unstable and selection of the final model was improved by resorting to 5-fold cross-validated lasso logit regression that has the capacity to improve out-of-sample predictions. Another study from Wuhan^9^ input data from 375 patients in a machine learning algorithm, and found that lactate dehydrogenase, lymphocyte count and CRP as the most significant predictors of severity, which was not confirmed in this study as lymphocyte count per se did not have any predictive value.

Another study from China examined 487 patients in Zhejiang Province to establish a score distinguishing high risk patients.^10^ They identified older age, male gender and presence of hypertension as predictors of severe disease at time of admission. We examined the effects of several comorbidities in our model including hypertension, ischemic heart disease, chronic respiratory disease, renal insufficiency and diabetes mellitus but found no predictive value over and above age which is consistent with recent data regarding the age component of fatality.^11^

A systematic review by Wynants et al identified 31 prediction models related to COVID19.^12^ The review included 10 prognostic models, all using data from China for predicting mortality risk, progression to severe disease, or length of hospital stay. The predictors included in more than one prognostic model from these studies were age (n=5), sex (n=2), features derived from CT scoring (n=5), C reactive protein (n=3), lactate dehydrogenase (n=3), and lymphocyte count. We again picked up several of these variables (except lactate dehydrogenase and lymphocyte count) when we ran standard regression modeling but, as explained previously, the model selection process was unstable when different randomly selected training samples were used. None of these studies based their model on consecutive patients or patients with a diagnosis of COVID19 not requiring hospital admission. One of the advantages of our study is that and because of regulations in Kuwait, all patients with PCR positivity confirming COVID19 are admitted to a hospital designated for COVID19. Kuwait enforced tough measures to identify and isolate patients with COVID19 since February 24^th^ when the first case was reported in the country from travelers to Iran. All patients coming from countries with COVID19 were quarantined for 2 weeks in quarantine institutions. There is currently a full border lockdown with partial curfew daily for 16 hours as well as closure of schools, Universities, government offices and businesses. These measures along with lockdown and aggressive testing in hot spots in Kuwait, allowed us to capture most patients with the disease in Kuwait which makes our data reported in this study truly representative of patients with COVID19 at time of diagnosis. Our predictive model is simple and most importantly determined at time of diagnosis/presentation which allows for distribution of resources and prioritization. With increase in cases with COVID19, the health system will not be able to afford to admit all patients even the asymptomatic ones to hospitals, and having such score that is reliable with regards to out-of sample predictions will allow stratification of patients for admission. Also, when more data is available regarding treatment for COVID-19, these prediction models can be used to identify patients at high risk to start treatment early. Therefore prediction models based on all patients with COVID-19 at the time of diagnosis, will serve the clinical purpose of utilizing rapidly diminishing resources better.

Our moderate to severe group of patients included those who died, were admitted to ICU, received ECMO, received supportive respiratory and renal treatment and stayed for more than 14 days in the hospital while either receiving treatment or demonstrating radiological signs or shortness of breath. This definition allowed us to capture the spectrum patients who truly required hospital support and treatment to allow prioritization of patients and their treatment at time of diagnosis

Although we internally validated our score, a limitation of this study is that it lacks external validation which we will be looking forward to perform with external institutions. Another limitation is the exclusion of patients not achieving the clinical course outcome of the study, including those with recent admission. Strengths include good discrimination and calibration results and use of a machine learning algorithm to improve out-of-sample predictions.

In conclusion, this simple prognostic score provides over-burdened health care systems during the pandemic with a much needed tool that can stratify patients at diagnosis. This should facilitate the decision making around admission versus home quarantine and will be of importance to the health care needs of the current pandemic.

## Data Availability

NA

## Supplementary material

### A. Calibration plots

1. In the validation cohort

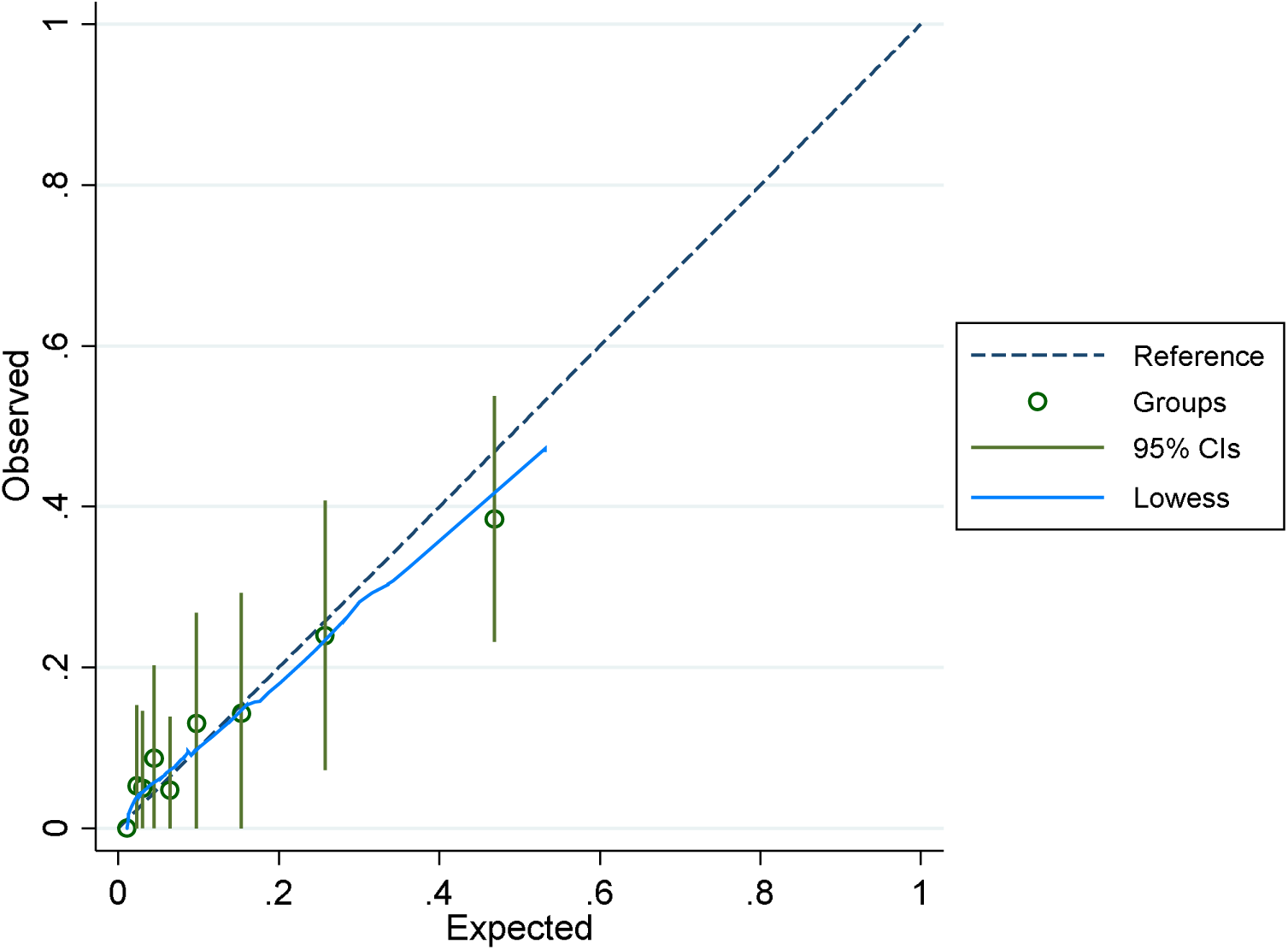
2. In the training cohort

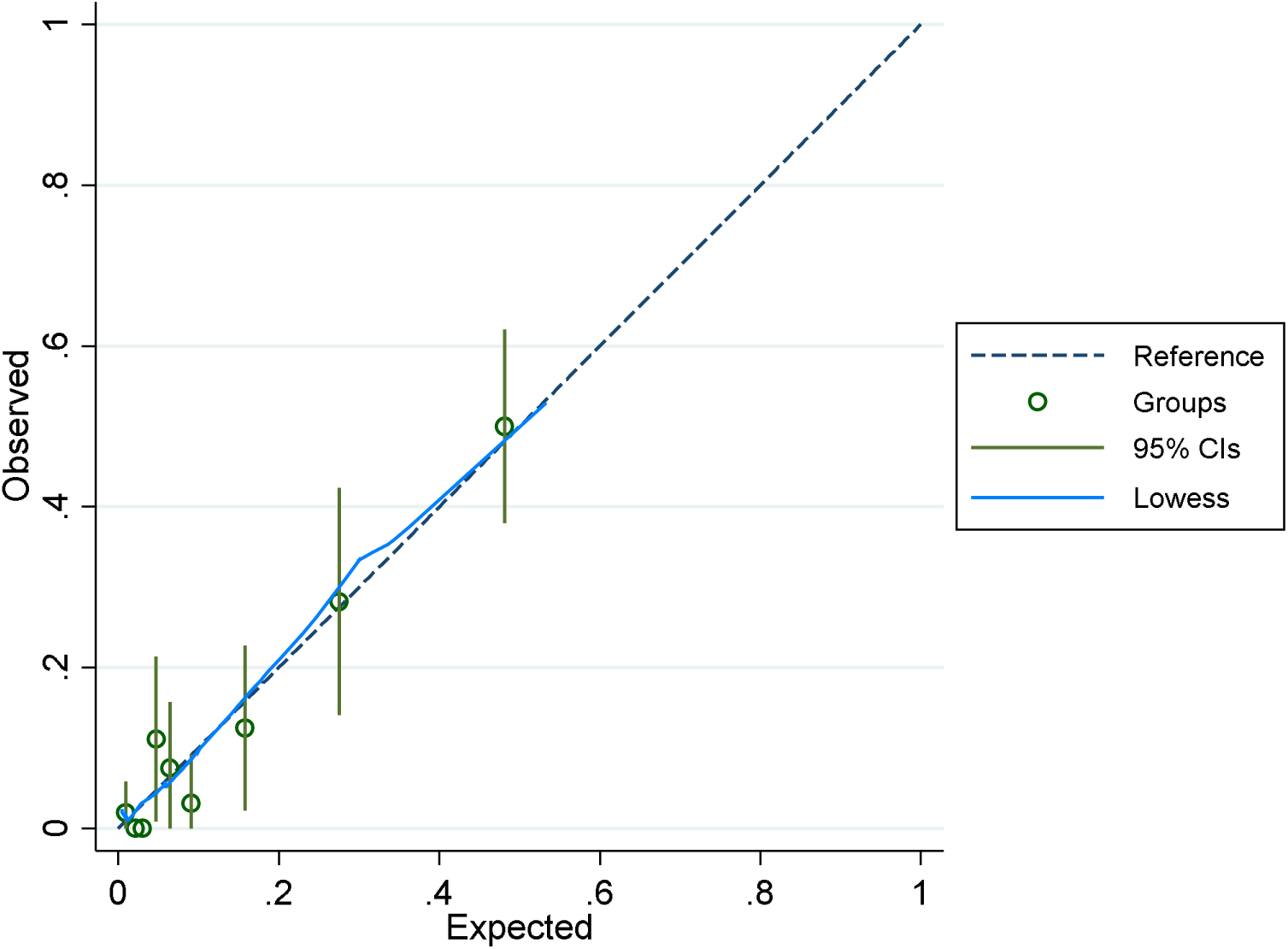

### B. Variables considered for the prediction model development

1. Measurements in blood: haemoglobin, platelet count, white cell count, lymphocyte%, monocyte%, red cell distribution width, INR, CRP, procalcitonin, creatinine, sodium, calcium, magnesium, albumin, urea, random glucose, Hba1c, gamma glutamyl transferase.
2. Symptoms: chills, cough, sputum production, sore throat, nasal congestion, conjunctival congestion, headache, fatigue, hemoptysis, nausea or vomiting, diarrhea, anosmia.
3. Comorbidities: diabetes mellitus, hypertension, coronary artery disease, chronic obstructive pulmonary disease, asthma, cerebrovascular disease, hepatitis, dyslipidemia, cancer, chronic renal disease, immunodeficiency.

### C. Test information

C-reactive protein (reference range 0- 8 mg/L) was measured by ELISA.

Procalcitonin (reference range 0.02-0.15 ng/ml) was measured by CMIA and CLIA.

Lymphocyte (reference range 19.4– 44.9%) and monocyte (reference range 0– 10.9%) percentages were determined by Complete Blood Count.

Albumin (reference range (35 – 48 g/L) was measured by biochemical, dye binding technique.

